# High fat and low carbohydrate supplies are linked to decreased epilepsy disease burden globally

**DOI:** 10.1101/2024.03.10.24304051

**Authors:** Duan Ni, Alistair Senior, David Raubenheimer, Stephen J. Simpson, Ralph Nanan

## Abstract

**Objectives:** Epilepsy is one of the major neural disorders globally. Ketogenic diets with high fat, low carbohydrate and moderate to low protein contents are well-established as interventions for epilepsy, particularly the intricate ones, exemplifying that modifying dietary compositions might have profound effects on established epilepsy. However, most of the diet-related epilepsy interventions have focused on dividual nutrients or specific diets with set nutrient compositions. An important unanswered question is whether specific macronutrient exposure through diets and food environments are linked to epilepsy and could potentially extend to primary preventive qualities.

**Methods:** Macronutrient supply, gross domestic product (GDP), and epilepsy disease burden data were collated from more than 150 countries spanning from 1990 to 2018. Nutritional geometry generalized additive mixed models (GAMMs) were carried out for analysis.

**Results:** GAMM modelling unravelled the interactive effects of nutrient supplies and socioeconomic status on epilepsy disease burden. Carbohydrate supply was associated with increased epilepsy while fat supply had the opposite effect. A high fat low carbohydrate supplies dietary environment, similar to ketogenic diets, was linked to the lowest epilepsy disease burden. These associations were conserved across sexes and were not confounded by the total energy supply.

**Conclusions:** A high fat low carbohydrate supplies dietary environment is associated with decreased epilepsy disease burden, hinting a plausible primary preventive role. This might expand the clinical application of ketogenic diets and inform future nutrient-based epilepsy treatment and/or prevention.

---

We read with interest the study by Qiao *et al*., “Ketogenic diet-produced β-hydroxybutyric acid accumulates brain GABA and increases GABA/glutamate ratio to inhibit epilepsy”^1^. Their study provides mechanistic insights into the well-established therapeutic effects of ketogenic diets for epilepsy^2, 3^.

This study exemplified that modifying dietary macronutrient compositions, like ketogenic diets with high fat, low carbohydrate contents, and moderate to low protein, might have profound effects on established epilepsy. However, the majority of diet-related epilepsy interventions have concentrated on individual macro- or micro-nutrients or specific diets with set nutrient compositions^3, 4^. An important unanswered question is whether specific macronutrient exposures via diets and food environments are linked to epilepsy, and thus could potentially extend to primary preventive qualities.

Here, harnessing nutritional geometry, a multi-dimensional analytical framework for disentangling relationships between dietary mixtures and outcomes like diseases, we analyzed global associations between nutrient supplies, a proxy for nutritional environments, and epilepsy disease burden. Importantly, our analyses used generalized additive mixed models (GAMMs), which account for interactions between nutrients, including their non-linear effects, which underly many facets of health and diseases, and adjust for potential confounding effects from socioeconomic wealth^5^.

Idiopathic epilepsy disease burden data was retrieved from the Global Burden of Disease database. Nutrient supply and global gross domestic product (GDP) data, as an indicator of the socioeconomic wealth, were obtained as previously described^6, 7^. An array of GAMMs were utilized for the analyses of the impacts from nutrient supplies and GDP on epilepsy disease burden globally over time.

As shown in Figure 1A, globally, epilepsy prevalence and incidence steadily increased from 1990 to 2018. This is accompanied with concurrent increases in global GDP and changing nutritional supply landscapes, with the most striking increase in fat supply (Figure 1B). Epilepsy incidence exhibits a heterogenous global distribution, affecting most countries. In 2018, there are 159 countries with data records, and the highest incidence was reported in Saudi Arabia, and the lowest was in Kiribati (Figure 1C).

**Figure 1.**
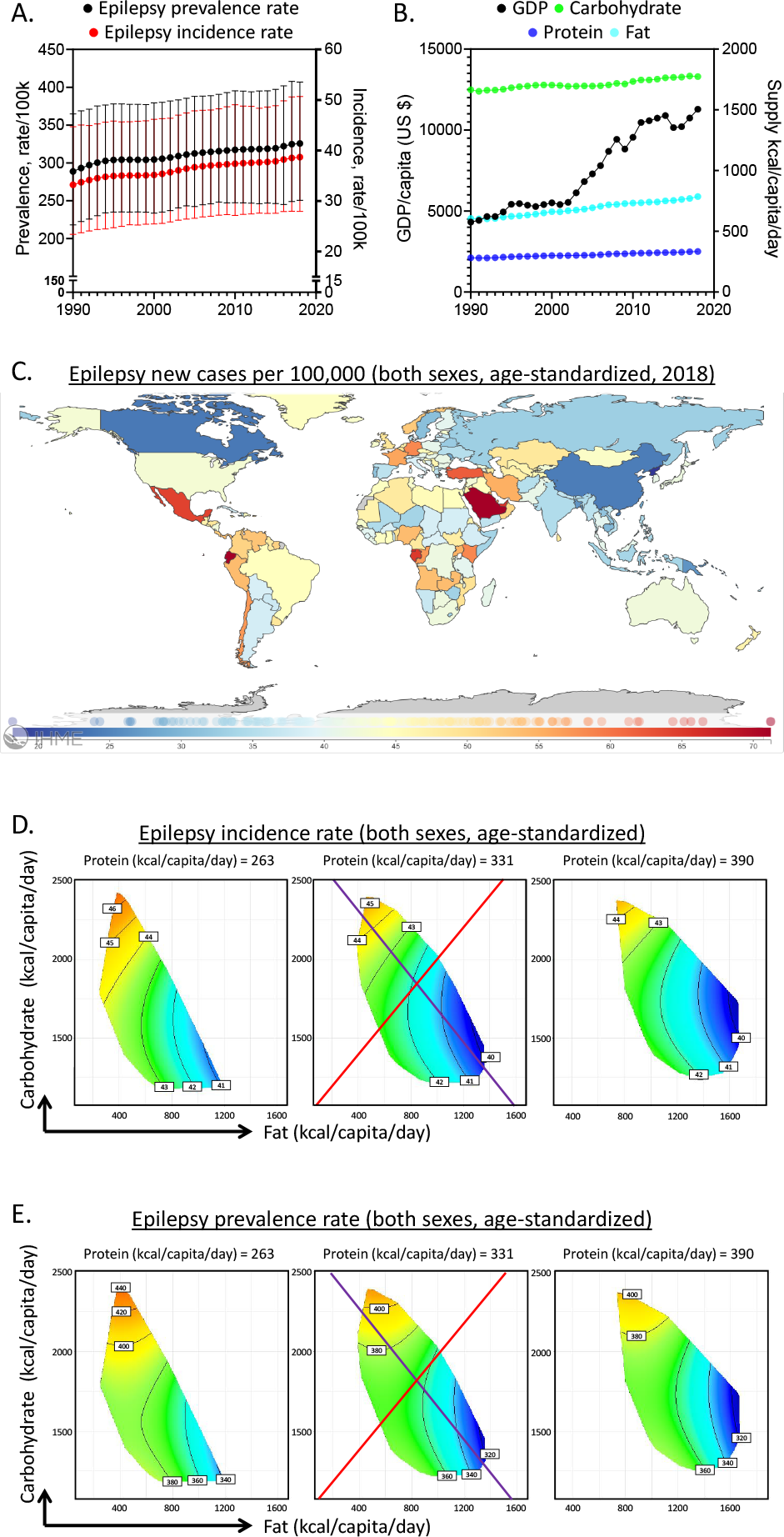
Global associations of macronutrient supplies and epilepsy disease burden. **A**. Global age-standardized prevalence (black) and incidence (red) of epilepsy of both sexes as functions of years. B. Global gross domestic product (GDP) per capita (in US dollars, black), and macronutrient supplies (carbohydrate: green, protein: blue, fat: cyan) as functions of years. **C**. A global overview of the age-standardized epilepsy incidence of both sexes in 2018. **D**. Modelled effects of macronutrient supplies on age-standardized epilepsy incidence rate of both sexes. **E**. Modelled effects of macronutrient supplies on age-standardized epilepsy prevalence rate of both sexes. (See Supplementary Information for statistics and interpretations).

To interrogate the potential associations between epilepsy disease burden and nutrient supplies, a series of GAMMs were explored, which included combinations of factors including nutrient-nutrient interactions, as well as changes over time and impacts from socioeconomic status. A model considering the interactions between macronutrient supplies and GDP, with an additive effect of time was favoured (Supplementary Table1, 3), illustrating the interactive effects of macronutrient supplies and socioeconomic status on epilepsy disease burden.

The results from 2018 are presented in Figure 1D as an example. This was the most recent year with relatively comprehensive data coverage. The modelled associations between macronutrient supplies and epilepsy incidence was presented as response surfaces mapped onto macronutrient supply plots. We focused on the effects of fat (*x*-axis) and carbohydrate (*y*-axis) supplies as they are the main variables in ketogenic diets. Protein supply was held at 25% (low), 50% (median) and 75% (high) quantiles of global supply. Within the modelling surfaces, red represented higher, while blue denoted lower epilepsy incidences.

Our modelling found that carbohydrate supply was strongly correlated with an increased epilepsy incidence, while fat supply had the opposite association, even after accounting for total energy supply (Figure 1D & Supplementary Table 1-2). This is visualized via the purple isocaloric line. Along this vector, the total nutrient energy supply was held constant, and carbohydrate was isocalorically replaced with fat. A higher fat:carbohydrate ratio was linked to lower epilepsy burden, with the lowest epilepsy incidence found in an environment with high fat but low carbohydrate supplies (bottom right), with macronutrient compositions similar to ketogenic diets. Across increasing quantiles of protein supplies, there was a mild change of epilepsy incidence, suggesting a less prominent impact from protein. Epilepsy incidence appeared less influenced by the total energy supply from macronutrients, because along the red radial, altering the total energy supply but keeping the fat:carbohydrate unchanged, minimally impacted epilepsy disease burden.

Similarly, carbohydrate supply was also associated with increased epilepsy prevalence while fat supply had the opposite effect (Figure 1E & Supplementary Table 3-4). These associations were independent of sex (Supplementary Figure S1A-B). Notably, further analyses by sub-dividing plant- and animal-based fats revealed that they conferred similar effects on epilepsy disease burden (Supplementary Figure S1C).

Collectively, our work represents the first study to correlate epilepsy disease burden with global nutritional environments, as reflected by nutrient supplies. Our analyses unveil a potential beneficial role of fat for epilepsy disease burden, after correcting its plausible interactions with other macronutrients, total energy supply and socioeconomic status. Increased fat supply is linked to reduced epilepsy disease burden, particularly coupled with decreased carbohydrate supply, reflecting a dietary environment similar to ketogenic diets.

As in the original paper and other studies, ketogenic diets used in epilepsy tend to be relatively low in protein. Our analyses show that protein supply only mildly influenced the epilepsy disease burden, requiring further research to probe how protein contents might modify the effects of ketogenic diets. This might also be relevant for improving the compliance of ketogenic regimens. On the other hand, whilst the therapeutic effects of ketogenic diets for recalcitrant epilepsy are well-established, their effects in reducing the general incidence of epilepsy are unknown. Our analyses based on nutrient supplies, which might only partly reflect the actual dietary and nutritional patterns of the populations on an individual level, indicate a possible primary preventive role of a high fat low carbohydrate nutrient environment against epilepsy. This warrants further explorations for both ketogenic diets and less extreme high fat, moderate to low protein and low carbohydrate diets. Also, it remains to be interrogated at which developmental stage exposure to a high fat low carbohydrate environment will be the most beneficial for epilepsy prevention. Moreover, the original study discovered that ketone body β-hydroxybutyric acid is the main contributor to the therapeutic effect of ketogenic diets in epilepsy. Therefore, it would also be of interest to inspect which threshold levels of ketosis are required for epilepsy treatment or prevention, which can potentially instruct the design of more stratified ketogenic dietary regimens for epilepsy.

The underlying mechanisms of the amelioration of epilepsy by ketogenic diets seem to be multifaceted, involving both the gut-brain axis and microbiota^8^, and regional changes within the brain including accumulation of gamma-aminobutyric acid (GABA) and elevated GABA/glutamate ratio^1^. Interestingly, reduction in GABA was also observed in attention-deficit/hyperactivity disorder (ADHD), another neuronal disorder that could potentially be alleviated by ketogenic diets^9^. Recently, using similar analyses, we found that a high fat low carbohydrate food environment is also associated with reduced ADHD disease burden^10^. This suggested that the mechanisms underlying the beneficial roles of ketogenic diets in epilepsy and ADHD might be partially shared, further hinting the therapeutic potentials of ketogenic diets in other GABA-related neurological or mental disorders.

Together, our ecological analyses have revealed a robust association between a nutrient environment with high fat and low carbohydrate supplies and decreased epilepsy disease burden. This might expand the clinical application of ketogenic diets and inform future nutrient-based epilepsy treatment and/or prevention.

## Data Availability

All data produced in the present study are available upon reasonable request to the authors

## Acknowledgements

This project is supported by the Norman Ernest Bequest Fund.

## Author Contributions

*Concept and design:* D.N., and R.N..

*Acquisition, analysis, and interpretation of data:* D.N., A.S., D.R., S.S., and R.N..

*Drafting of the manuscript:* D.N., and R.N..

*Critical revision of the manuscript for important intellectual content:* All authors.

