## Supplementary Information for "High fat and low carbohydrate supplies are linked to decreased epilepsy disease burden globally"

### Interpretation of modelling surfaces

Details for modelling interpretation can be referred to previous publications[1-3]. In brief, using RStudio (v4.2.2.), epilepsy disease burden data were analyzed utilizing generalized additive mixed models (GAMMs)[4, 5]. Results from analysis of the supplies of three macronutrients were shown as response surfaces based on nutrient axes for three macronutrients, protein, carbohydrate and fat. For analysis of different fat types, results were plotted on response surfaces with plant-based fat as the $x$-axis and animal-based fat as the $y$-axis. The sums of carbohydrate and protein supplies were held at 25%, 50% and 75% of the global data.

Within the response surfaces, red means higher values, and blue implies lower. Along the black contour lines, the modelled values are constant and the numbers on them denote the magnitude of the parameters. The purple line is an isocaloric vector, along which the total energy supply from macronutrients is unchanged but fat is isocalorically substituted with carbohydrate. The food rail is depicted by the red line. Carbohydrate:fat ratio is held constant along it, while the total energy supply is altered.

Statistics for the modelling analysis are shown in Supplementary Tables. When the modelling analyses are significant, effects of macronutrient supplies on the modelled values (i.e., disease burden) can be deduced based on the modelling surfaces.

### Supplementary Methods

#### *Data collection and processing*

Data collation and processing followed the workflow described previously[1, 3]. Epilepsy data was extracted from the Global Burden of Disease Study 2019. As described[1], macronutrient supply and gross domestic product (GDP) data were from the Food and Agriculture Organization Corporate Statistical Database (FAOSTAT, www.fao.org/faostat/en/#home) and the Maddison project[6] respectively.

Countries or time points with no data record were excluded and the resulting data spanning from 1990 to 2018 covering more than 150 countries, covering all continents, were further analyzed with R.

*Generalized additive mixed models (GAMMs)*

Details of the models were described in[1, 3]. In brief, GAMMs[3,4] were run to model the changes of epilepsy burden over time and disentangle the influences from macronutrient supply and GDP. Overall, GAMMs share similar assumptions to generalized linear models. They account for the nonlinear terms as nonparametric smoothed functions, often in a form of spline, and provide a flexible manner to estimate the nonlinear associations. All analyses were run using the *mgcv* package and its "gam" function[4, 5]. All models took into account the countries that the data were from as random effects. The gamma parameter, implying the smoothing degrees of the modelled effects, was calculated as $\log(n)/2$, and $n$ is the number of combinations for countries and years with our dataset. A Gaussian family with log-link function was used for modelling.

A series of different predictor variables and their different combinations and a null model where only the random effects from countries are considered are compared. Models with multiple variables consider all combinations of the individual, additive and interactions among parameters like macronutrient supply, year and GDP data. Macronutrient supply was modelled as a three-dimensional spline, and year and GDP data were modelled as one-dimensional cubic-regression splines utilizing the "s()" function in *mgcv* package. The interactions between smooth terms on different scales such as macronutrient supply and year were modelled with "te()" function from *mgcv* package using tensor product smoothers.

Modelling results were evaluated using Akaike information criterions (AICs) and the model with the lowest AIC was favoured[7].

Codes for the analysis are adapted from GitHub, https://github.com/Nidane/Asthma-NutrientSupply.

**Supplementary Tables**

Supplementary Tables 1-4 are GAMMs estimates. For parametric terms the model estimates and associated standard errors (SE) and test statistics are presented. For non-parametric smooth terms, the estimated and reference degrees of freedom (reflected by edf, sumEDF, Ref.df) are shown as well as their test statistics. Smooth terms were fitter with either the standard smooth function “s()” or a tensor product smooth “te()” in the *mgcv* package. Tensor product smoothing was utilized where terms exist on different units (for example, nutrient supply and year). Country was included as a random effect using the smooth function “s()”. The model family and implemented link function is stated.

**Supplementary Table 1.** Relative fit of generalized additive mixed models (GAMMs) testing the predictors for age-standardized epilepsy incidence rate in both sexes. Gamma is the degrees of freedom inflation factor. Dev means the deviance explained. AIC = Akaike information criterion. GDP = gross domestic product. Delta is the differences between AICs of models and the minimum AIC. sumEDF reflects the degrees of freedom of the models. Macronutrient supply was modelled as a three-dimensional thin-plate spline. (related to Figure 1D)

| GAM | Gamma | Dev | AIC | Delta | Weights | sumEDF | Formula |
|---|---|---|---|---|---|---|---|
| 1 | 3.855439 | 94.9937 | 19265.04 | 1582.696 | 0 | 156.7595 | 1 + s(Country, bs="re") |
| 2 | 3.855439 | 95.57433 | 18733.33 | 1050.987 | 6.04046520236329e-229 | 166.1167 | s(protein.kcal, carb.kcal, fat.kcal, k=k_nut) + s(Country, bs="re") |
| 3 | 3.855439 | 95.82611 | 18461.05 | 778.7025 | 8.07034287889806e-170 | 160.737 | s(Year, k=10, bs="cr") + s(Country, bs="re") |
| 4 | 3.855439 | 96.12953 | 18134.76 | 452.4134 | 5.75010450512984e-99 | 166.0875 | s(GDP, k=10, bs="cr") + s(Country, bs="re") |
| 5 | 3.855439 | 95.99348 | 18297.16 | 614.8217 | 3.11291746915237e-134 | 170.1643 | s(protein.kcal, carb.kcal, fat.kcal, k=k_nut) + s(Year, k=10, bs="cr") + s(Country, bs="re") |
| 6 | 3.855439 | 96.22109 | 18046.3 | 363.9575 | 9.28203611830669e-80 | 175.306 | s(protein.kcal, carb.kcal, fat.kcal, k=k_nut) + s(GDP, k=10, bs="cr") + s(Country, bs="re") |
| 7 | 3.855439 | 96.2293 | 18025.59 | 343.2509 | 2.91087367437179e-75 | 169.8055 | s(Year, k=10, bs="cr") + s(GDP, k=10, bs="cr") + s(Country, bs="re") |
| 8 | 3.855439 | 96.17783 | 18101.43 | 419.0882 | 9.91204695567263e-92 | 177.4561 | te(protein.kcal, carb.kcal, fat.kcal, Year, bs=c("tp", "cr"), d=c(3,1), k=c(k_nut, 7)) + s(Country, bs="re") |
| 9 | 3.855439 | 96.48861 | 17763.12 | 80.77542 | 2.88297907713333e-18 | 197.6296 | te(protein.kcal, carb.kcal, fat.kcal, GDP, bs=c("tp", "cr"), d=c(3,1), k=c(k_nut, 7)) + s(Country, bs="re") |
| 10 | 3.855439 | 96.27368 | 17983.38 | 301.0401 | 4.26553916109995e-66 | 175.1363 | te(Year, GDP, k=10) + s(Country, bs="re") |
| 11 | 3.855439 | 96.46425 | 17772.05 | 89.71024 | 3.30878117812915e-20 | 186.6639 | te(protein.kcal, carb.kcal, fat.kcal, Year, bs=c("tp", "cr"), d=c(3,1), k=c(k_nut, 7)) + s(GDP, k=10, bs="cr") + s(Country, bs="re") |
| **12** | **3.855439** | **96.55677** | **17682.34** | **0** | **1** | **201.0077** | **te(protein.kcal, carb.kcal, fat.kcal, GDP, bs=c("tp", "cr"), d=c(3,1), k=c(k_nut, 7)) + s(Year, k=10, bs="cr") + s(Country, bs="re")** |
| 13 | 3.855439 | 96.35707 | 17900.46 | 218.1142 | 4.33611343332191e-48 | 184.1969 | te(Year, GDP, k=10) + s(protein.kcal, carb.kcal, fat.kcal, k=k_nut) + s(Country, bs="re") |

**Supplementary Table 2.** Estimated effects of macronutrient supply by time and GDP per capita on age-standardized epilepsy incidence rate. Gaussian-GAMM, log-link function. (related to Figure 1D)

| **Parametric coefficients** | | | | |
|---|---|---|---|---|
| | **Estimate** | **Std. Error** | **t value** | **Pr(>\|t\|)** |
| **(Intercept)** | 3.71294 | 0.01113 | 333.7 | <2e-16 |
| **Approximate significance of smooth terms** | | | | |
| | **edf** | **Ref.df** | **F** | **p-value** |
| **te(protein.kcal,carb.kcal,fat.kcal,GDP)** | 39.886 | 44.592 | 18.38 | <2e-16 |
| **s(Year)** | 4.096 | 5.048 | 18.56 | <2e-16 |
| **s(Country)** | 157.026 | 158.000 | 498.62 | <2e-16 |
| R-sq.(adj) = 0.964 Deviance explained = 96.6% | | | | |
| GCV = 4.1155 Scale est. = 2.9379 n = 4465 | | | | |

**Supplementary Table 3.** Relative fit of generalized additive mixed models (GAMMs) testing the predictors for age-standardized epilepsy prevalence rate in both sexes. Gamma is the degrees of freedom inflation factor. Dev means the deviance explained. AIC = Akaike information criterion. GDP = gross domestic product. Delta is the differences between AICs of models and the minimum AIC. sumEDF reflects the degrees of freedom of the models. Macronutrient supply was modelled as a three dimensional thin-plate spline. (related to Figure 1E)

| GAM | Gamma | Dev | AIC | Delta | Weights | sumEDF | Formula |
|---|---|---|---|---|---|---|---|
| 1 | 3.855439 | 94.99174 | 41350.68 | 1160.188 | 1.17032895065262e-252 | 156.5077 | 1 + s(Country, bs="re") |
| 2 | 3.855439 | 95.24388 | 41138.64 | 948.1556 | 1.29013552243245e-206 | 165.8156 | s(protein.kcal, carb.kcal, fat.kcal, k=k_nut) + s(Country, bs="re") |
| 3 | 3.855439 | 95.18793 | 41180.49 | 990.0003 | 1.0572038061995e-215 | 160.6266 | s(Year, k=10, bs="cr") + s(Country, bs="re") |
| 4 | 3.855439 | 95.6942 | 40694.67 | 504.1767 | 3.3069004380407e-110 | 165.8899 | s(GDP, k=10, bs="cr") + s(Country, bs="re") |
| 5 | 3.855439 | 95.38353 | 41013.33 | 822.8458 | 2.09557225809927e-179 | 169.6911 | s(protein.kcal, carb.kcal, fat.kcal, k=k_nut) + s(Year, k=10, bs="cr") + s(Country, bs="re") |
| 6 | 3.855439 | 95.81011 | 40591.04 | 400.5542 | 1.04895520077913e-87 | 174.9974 | s(protein.kcal, carb.kcal, fat.kcal, k=k_nut) + s(GDP, k=10, bs="cr") + s(Country, bs="re") |
| 7 | 3.855439 | 95.80002 | 40592.82 | 402.3344 | 4.30727571856498e-88 | 170.5212 | s(Year, k=10, bs="cr") + s(GDP, k=10, bs="cr") + s(Country, bs="re") |
| 8 | 3.855439 | 95.70236 | 40728.9 | 538.4163 | 1.21443805985666e-117 | 187.2449 | te(protein.kcal, carb.kcal, fat.kcal, Year, bs=c("tp", "cr"), d=c(3,1), k=c(k_nut, 7)) + s(Country, bs="re") |
| 9 | 3.855439 | 96.11332 | 40305.23 | 114.7441 | 1.2123890193063e-25 | 199.7983 | te(protein.kcal, carb.kcal, fat.kcal, GDP, bs=c("tp", "cr"), d=c(3,1), k=c(k_nut, 7)) + s(Country, bs="re") |
| 10 | 3.855439 | 96.00499 | 40403.92 | 213.4333 | 4.50337432946742e-47 | 187.7718 | te(Year, GDP, k=10) + s(Country, bs="re") |
| 11 | 3.855439 | 96.14204 | 40271.01 | 80.5263 | 3.2654037777047e-18 | 199.2484 | te(protein.kcal, carb.kcal, fat.kcal, Year, bs=c("tp", "cr"), d=c(3,1), k=c(k_nut, 7)) + s(GDP, k=10, bs="cr") + s(Country, bs="re") |
| **12** | **3.855439** | **96.22044** | **40190.49** | **0** | **1** | **204.818** | **te(protein.kcal, carb.kcal, fat.kcal, GDP, bs=c("tp", "cr"), d=c(3,1), k=c(k_nut, 7)) + s(Year, k=10, bs="cr") + s(Country, bs="re")** |
| 13 | 3.855439 | 96.06201 | 40354.17 | 163.6792 | 2.86749926292708e-36 | 194.9879 | te(Year, GDP, k=10) + s(protein.kcal, carb.kcal, fat.kcal, k=k_nut) + s(Country, bs="re") |

**Supplementary Table 4.** Estimated effects of macronutrient supply by time and GDP per capita on age-standardized epilepsy prevalence rate. Gaussian-GAMM, log-link function. (related to Figure 1E)

| **Parametric coefficients** | | | | |
|---|---|---|---|---|
| | **Estimate** | **Std. Error** | **t value** | **Pr(>\|t\|)** |
| **(Intercept)** | 5.91261 | 0.02423 | 244 | <2e-16 |
| **Approximate significance of smooth terms** | | | | |
| | **edf** | **Ref.df** | **F** | **p-value** |
| **te(pbf.kcal,carb_prot.kcal,abf.kcal,GDP)** | 42.963 | 48.294 | 21.67 | <2e-16 |
| **s(Year)** | 4.275 | 5.258 | 17.45 | <2e-16 |
| **s(Country)** | 157.580 | 158.000 | 458.18 | <2e-16 |
| R-sq.(adj) = 0.960 Deviance explained = 96.2% | | | | |
| GCV = 640.38 Scale est. = 453.91 n = 4465 | | | | |

**Supplementary Figure**

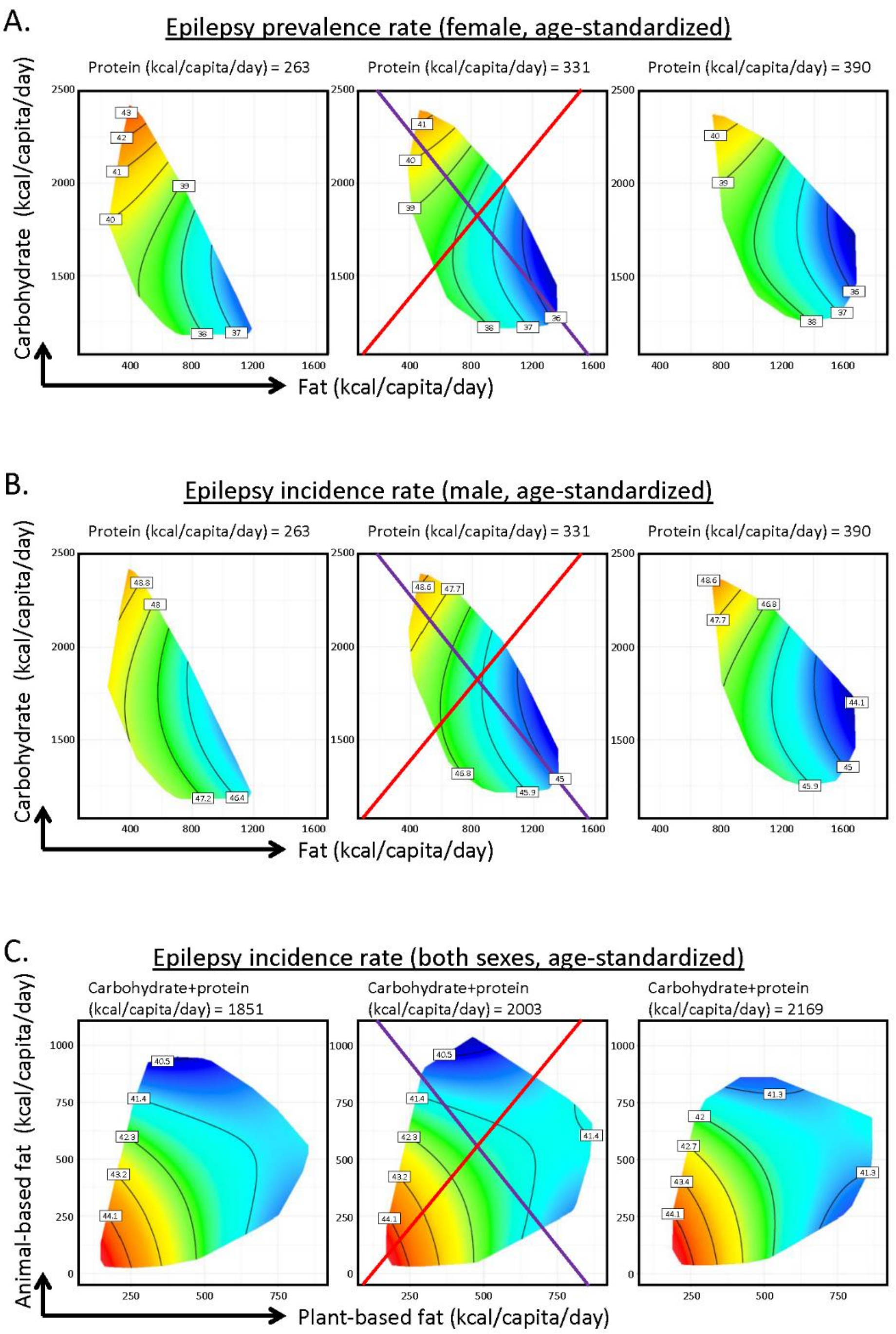


**Supplementary Figure S1.** Predicted effects of macronutrient supply on age-standardized epilepsy incidence rate of female (**A**), age-standardized epilepsy incidence rate of male (**B**) and predicted effects of plant-based versus animal-based fat supply on age-standardized epilepsy incidence of both sexes (**C**).
